# Evaluation of post-acute COVID-19 health outcomes (ECHOES) in England: The development of national surveillance system for long-term health outcomes following COVID-19

**DOI:** 10.1101/2024.10.18.24315744

**Authors:** Hester Allen, Katie Hassell, Christopher Rawlinson, Owen Pullen, Colin Campbell, Annika M. Jödicke, Martí Català, Albert Prats Uribe, Gavin Dabrera, Daniel Prieto-Alhambra, Ines Campos-Matos

**Author notes:** **Correspondence:** Hester Allen.

## Abstract

**Introduction:** Electronic health records can be used to understand the diverse presentation of post-acute and long-term health outcomes following COVID-19 infection. In England, the UK Health Security Agency in collaboration with the University of Oxford have created the ECHOES dataset to monitor how an initial SARS-CoV-2 infection episode is associated with changes in the risk of health outcomes that are recorded in routinely collected health data.

**Methods:** The ECHOES dataset is as a national level dataset combining national level surveillance, administrative, and healthcare data. Entity resolution and data linkages methods are used to create a cohort of individuals who have tested positive and negative for SARS-CoV-2 in England throughout the COVID-19 pandemic, alongside information on a range of health outcomes including diagnosed clinical conditions and mortality, and risk factor information.

**Results:** The dataset contains comprehensive COVID-19 testing data and demographic, socio economic and health related information for 44 million individuals, who tested for SARS-CoV-2 between March 2020 and April 2022, representing 15,720,286 individuals who tested positive and 42,351,016 individuals who tested negative.

**Discussion:** With the application of epidemiological and statistical methods, this dataset allows a range of clinical outcomes to be investigated, including pre-specified health conditions and mortality. Furthermore, understanding of potential determinants of health outcomes can be gained, including pre-existing health conditions, acute disease characteristics, SARS-CoV-2 vaccination status and genomic variant.

## Introduction

SARS-CoV-2 infection typically causes an acute respiratory syndrome, and there has been significant research into different health outcomes within the acute period following symptom onset, such as critical care admission and death.

Increasingly, data driven approaches to characterise and determine the prevalence, symptoms and risk factors of post-acute COVID-19 are being developed (4, 5). Electronic health records can be used to understand the diverse presentation of post-acute and long-term COVID-19 heath outcomes in representative populations and in routine care (3).

In England, during the COVID-19 pandemic, robust, national surveillance systems were developed to collect comprehensive data to monitor trends in COVID-19. Using these data, UKHSA, in collaboration with the University of Oxford, have developed the Evaluation of Post-acute COVID-19 Health Outcomes (ECHOES) dataset.

The purpose of this surveillance dataset is to monitor how an initial SARS-CoV-2 infection episode is associated with changes in the risk of health outcomes that are recorded through secondary care use, and death.

To overcome limitations of other publications in the literature, of which many have focused on those with the most severe acute SARS-CoV-2 infection, we developed this dataset to include a broad range of both community based and hospitalised cases, as well as a developed control group based on negative laboratory tests.

ECHOES is a dataset designed as a longitudinal cohort comprising individuals tested for SARS-CoV-2 in England throughout the COVID-19 pandemic. This dataset benefits from the use of a range of large data including, comprehensive, national level SARS-CoV-2 testing data, genomic sequencing data and vaccination records, alongside extensive demographic, socio economic and health related information.

The use of routine health record datasets allows health outcomes, including those presented in secondary healthcare settings to be assessed using pre-existing methodologies, and therefore outcomes can be comparable during the exposure and follow-up periods and between persons with SARS-CoV-2 infection and test negative controls.

As part of the remit to diagnose, control, prevent, and describe trends in communicable diseases and other risks to public health, UKHSA processes personal information to develop and maintain surveillance systems and conduct infection control activities(7), in accordance with Regulation 3 SI 1438 Control of Healthcare Information Regulations 2002 (s251)(8) as the legal basis for data processing in the absence of individual consent.

## Methods

The ECHOES dataset is a longitudinal cohort capturing individuals who tested for SARS-CoV-2 between March 2020 and May 2022. This cohort is linked to multiple data sources to obtain information on health outcomes occurring in the post-acute phase (defined as >28 days after test), and factors associated with the COVID-19 infection and health outcomes. Building of the dataset occurs in three steps: cohort building, linkage to additional data sources, and matching. Data processing is carried out using SQL and statistical analysis is carried out using R, working within UKHSA secure servers.

### Data sources

The data sources utilised to build the ECHOES dataset are all routinely collected national level surveillance, administrative, and healthcare data for which UKHSA are the owners or processors.

This dataset is built linking: COVID-19 testing data; the National Immunisation Management system (NIMS); the Office of National statistics All-cause mortality data; and Hospital Episode statistics.

COVID-19 testing data are collected through two data systems – the Second-Generation Surveillance System (SGSS) and the Unified Sample Dataset (USD).

***Second generation surveillance system (SGSS)*** collects infectious disease laboratory test information from diagnostic laboratories in England (4). SARS-CoV-2 testing data in SGSS includes polymerase chain reaction (PCR) tests from NHS settings (including NHS patients and staff) and PCR and lateral flow device (LFD) tests from community settings, submitted by individuals via an NHS website.

SGSS records are validated and enhanced using the NHS spine, a database containing person identifying information (PII) for everyone who has ever had a contact with the NHS(5). Positive test records are converted to cases and infection episodes, deduplicating records using the Organism-Patient-Illness Episode (OPIE) principle, whereby episodes constitute a positive organism in a defined time-period. For COVID-19, episodes are defined as 91 days from an individual’s earliest positive specimen date within a previous episode(4).

Where available, SARS-CoV-2 genomic information is included in SGSS(6). In England, during the pandemic, genomic investigation and variant assignment was coordinated by the COVID-19 Genomics UK consortium (COG UK). A sample of eligible SARS-CoV-2 PCR samples underwent genomic investigation via: i) whole genome sequencing ii) reflex (genotyping) assays to detect key mutations, and iii) S-gene target status of PCR tests carried out on the TaqPath assay.

***The Unified sample dataset (USD)*** is a repository for positive and negative COVID-19 testing data in England, including all PCR and LFD tests (7). Testing data are sourced from SGSS, Respiratory Datamart (a laboratory reporting system designed for influenza test data collection)(8) and the National Pathology Exchange (NPEX). Test records are enriched with patient information from the NHS spine (9).

***The national immunisation management system (NIMS)*** collects information on individuals eligible for SARS-CoV-2 vaccination and on administered vaccinations across a range of settings. It was commissioned in 2020 as part of the national influenza and COVID-19 vaccination programmes (13,14).

NIMS contains two data views: person records for all individuals issued with an NHS number in England (the NIMS population denominator) who were alive when the vaccination programme began in December 2020, identifying those eligible for vaccination; and detailed vaccination records for all individuals who have received at least one dose of SARS-CoV-2 vaccination.

Patient information is obtained from the NHS spine and undergoes validation to improve accuracy and completeness.

***All-cause mortality data*** are obtained from death registrations for England and Wales collated by the Office for National Statistics (ONS)(15). Data are reported at person level and contain information about the death and the deceased individual. Causes of death, including underlying and primary cause of death are provided as International Classification of Diseases (ICD-10) codes, obtained through using automatic coding software to convert text terms from the death certificate. Primary cause of death refers to the disease or condition directly leading to death and underlying cause is the significant conditions contributing to the death (16).

***Hospital Episode Statistics*** collects information about all NHS hospital admissions, Accident and Emergency (A&E) attendances and outpatient visits in England (17).

Standardised codes for procedures (Office of Population Censuses and Surveys (OPCS) codes) and diagnoses and medical conditions (ICD-10 codes), are captured for each episode, including information on primary diagnosis, treatment, co-morbidities and complications (20).

Supplementary Table 1 describes the features of the datasets used in the production of these data.

### Cohort building methods

To create a cohort of individuals who tested positive and negative for SARS-CoV-2 we identify unique individuals who have tested positive and negative within the COVID-19 testing datasets. This is done by applying entity resolution methods, which utilise PII and demographic information to create a dataset containing key identifiers for linkage to other data sources.

Entity resolution involves SGSS, USD, the NIMS denominator and death registration data. Individuals can be captured in both the SGSS and USD testing data multiple times. The entity resolution methods use linkage methods between these data and the NIMS and death registration data to identity unique individuals within the testing data. To date, COVID-19 test and death registration records from January 2020 to December 2023, and NIMS records for those eligible for vaccination from December 2020 to May 2022 are included in the entity resolution process.

To maximise uniformity, datasets are initially cleaned, removing all non-alphanumeric characters and additional spaces present in name, NHS number and postcode variables, and NHS numbers and postcodes are validated (9). The NIMS denominator acts as the gold-standard for PII as information is obtained from the NHS spine, and death registration data provides validated PII for deceased individuals.

The entity resolution process runs, firstly between SGSS, USD and NIMS and secondly between SGSS, USD and death registration data. Each stage consists of 5 sequential iterations (Table 1) and nine sub-iterations (Table 2). Once linked, individuals are excluded from following iterations. All iterations include deterministic matching of at least three person identifiers (of NHS number, sex, residential postcode and date of birth) and deterministic matching and probabilistic matching (using the LIKE and DIFF SQL functions) of forename and surname.

**Table 1:**
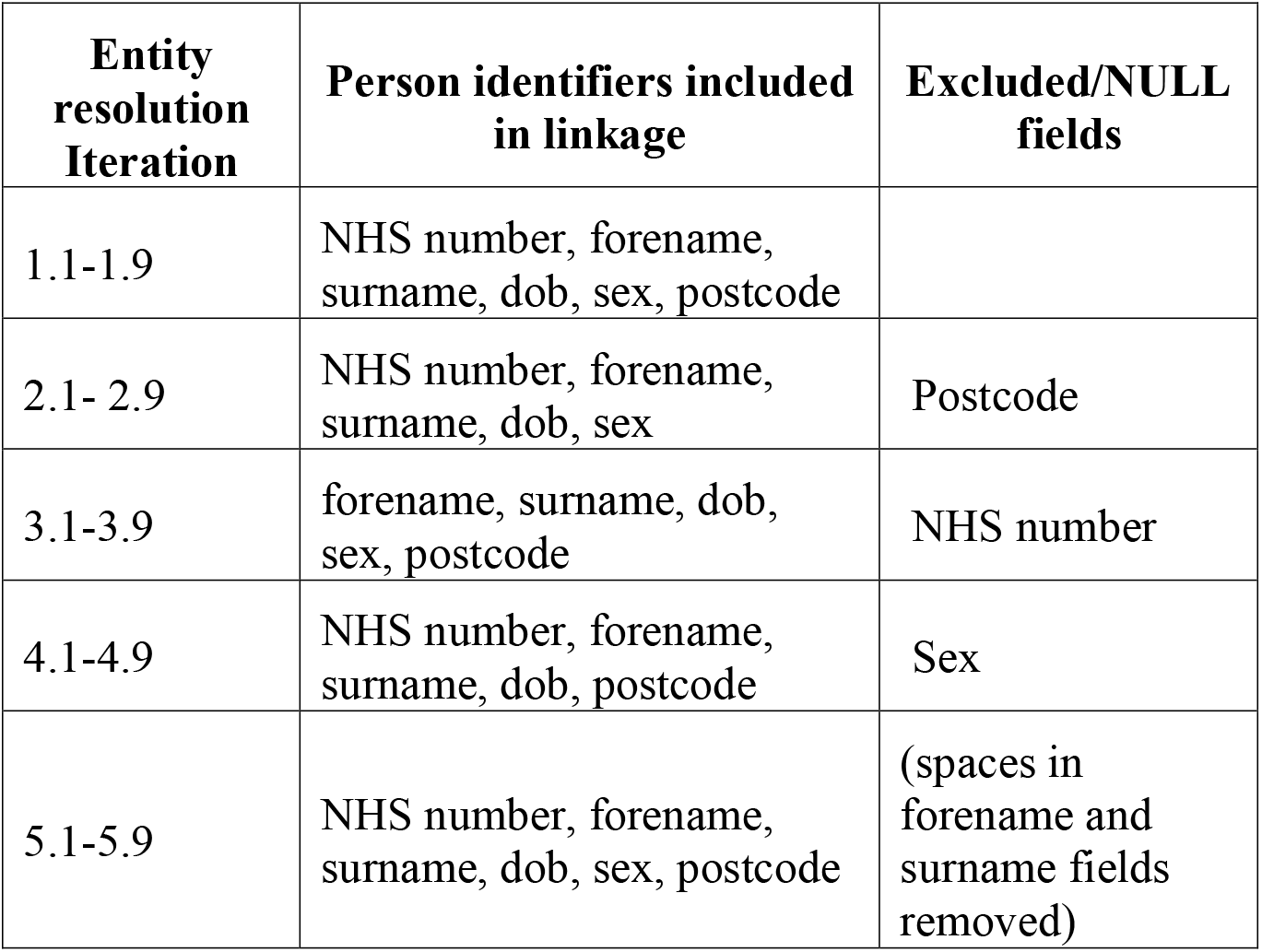
Description of entity resolution iterations 1.1 to 5.9 used in ECHOES cohort creation.

| **Entity resolution Iteration** | **Person identifiers included in linkage** | **Excluded/NULL fields** |
| --- | --- | --- |
| 1.1-1.9 | NHS number, forename, surname, dob, sex, postcode |  |
| 2.1- 2.9 | NHS number, forename, surname, dob, sex | Postcode |
| 3.1-3.9 | forename, surname, dob, sex, postcode | NHS number |
| 4.1-4.9 | NHS number, forename, surname, dob, postcode | Sex |
| 5.1-5.9 | NHS number, forename, surname, dob, sex, postcode | (spaces in forename and surname fields removed) |

**Table 2:** Description of entity resolution sub iterations 1 to 9 used in ECHOES cohort creation.

|  | **Person identifiers deterministically linked** | **Person identifiers probabilistically linked** |
| --- | --- | --- |
| 1 | NHS number, forename, surname, dob, sex, postcode |  |
| 2 | NHS number, surname, dob, sex, postcode | forename (LIKE*) |
| 3 | NHS number, surname, dob, sex, postcode | forename (DIFF**) |
| 4 | NHS number, forename, dob, sex, postcode | surname (LIKE) |
| 5 | NHS number, forename, dob, sex, postcode | surname (DIFF) |
| 6 | NHS number, dob, sex, postcode | forename (LIKE), surname (LIKE) |
| 7 | NHS number, dob, sex, postcode | forename (DIFF), surname (LIKE) |
| 8 | NHS number, dob, sex, postcode | forename (LIKE), surname (DIFF) |
| 9 | NHS number, dob, sex, postcode | forename (DIFF), surname (DIFF) |

\* The LIKE function links two records if a specific character string (such as “Chris”) matches a specified character pattern (such as “Christopher”)(10).
\*\*The DIFF function uses SOUNDEX to compare the difference between two character expressions (11). SOUNDEX converts string variables to four-character codes based on how the string sounds when spoken in English and can then be compared to other strings in terms of how they sound when spoken (12). For example, “Chris” would be linked to “Kris”.

Iteration one includes all identifiers (NHS number, forename, surname, DOB, sex and postcode). Subsequent iterations exclude postcode, NHS number and sex and consider alternative formatting of person identifiers. This account for where people may have changed residence over the time-period captured, where different formatting of names have been used across datasets (eg. Anne-Marie vs Anne Marie) and where key fields are missing, such as NHS number and erroneously entered, such as sex.

These iterations are carried out between the two testing data sources and NIMS denominator data, then for records not successfully linked, between testing data and ONS death registration data.

Following entity resolution, all PII except NHS number which is required for linkage to other data sources are removed from the dataset.

### 2.3 Censoring and exclusion criteria

Exclusion criteria and censoring rules are then applied. All records require a valid NHS number which has been verified against or obtained from NIMS or death registration data, therefore testing records not linked during the entity resolution process were excluded.

To investigate post-acute outcomes, individuals are required to have survived beyond the acute phase of infection, with follow-up starting 28 days after individuals’ earliest positive and negative tests dates. Data are at individual and test result level, as individuals can contribute both positive and negative follow-up time. If an individual tests negative before their first positive, both tests are included, however, if the positive test is within 28 days of the negative test, the negative test is negated, and the positive test retained. Positive test records are not negated if a negative test is recorded within 28 days.

To assess time at risk, follow-up time is calculated. For negatives, follow-up time is calculated from 28 days after their negative test, and whichever comes first of a) date of death, b) a positive test or, c) the health outcome of interest occurs. For positives, between earliest test date and date of death or outcome occurrence (Figure 2).

### Linkage to other data sources

The ECHOES cohort is linked (using NHS number) to other data sources to obtain information a range of health outcomes and factors associated with the SARS-CoV-2 infection and health outcomes. These include cause of death, pre-existing health conditions, comorbidities, and acute phase hospitalisation (Supplementary Table 1).

All-cause mortality provides date and cause of death. Time to death is calculated and cause of death ICD-10 codes are categorised into disease groups(13).

Vaccination status at time of test is obtained from NIMS and is defined in several ways: i) as fully vaccinated (having two or more vaccine doses) or unvaccinated, ii) by time since last vaccine dose received.

Linkage of the ECHOES cohort to HES allows a range of health indicators including acute disease characteristics and pre-existing health conditions to be developed, as well as post-acute health outcomes to be defined. Hospital admission information at the time of test can be used to indicate hospitalisation during the acute phase of COVID-19 as well as whether an individual’s SARS-CoV-2 test was carried out in a hospital setting. Diagnosis ICD-10 codes captured in HES can be used to indicate pre-existing conditions and comorbidities if recorded prior individual’s test. Specific pre-existing conditions can be defined and categorised using the standard ICD-10 chapter categorisations or through clinical phenotyping of diagnosis codes, and comorbidities scores can be assigned using tools such as the Charlson Comorbidity index(14). Furthermore, information on hospital admissions and procedures occurring during the post-acute phase can indicate healthcare usage, and by applying using similar disease area and condition categorisation methods, diagnosis codes during this phase can be used to define post-acute health outcomes within varying time periods after an individual’s initial SARS-CoV-2 test.

Following data linkage, all PII are removed from the dataset.

### Analytical method

The aim of analyses using these data is to compare the risk of post-acute health outcomes between those who tested positive and negative for SARS-CoV-2.

Using this cohort, a range of study designs can be used to assess different outcomes within individual’s follow-up time. A matched cohort design can be applied, matching individuals who test positive and negative on a range of characteristics, depending on the objectives of the study.

## Results

The entity resolution process using 16,598,060 positive episodes and 421,875,882 negative tests recorded at the time of dataset development result in 92.8% of positive records and 85.7% of negative records who tested between March 2020 and May 2022 being linked to a person record. Following censoring and exclusion, 44,234,762 unique individuals who have tested for COVID-19 are included. As individuals can be included in the positive and negative groups, this represents 15,720,286 positive individuals and 42,351,016 negative individuals (Figure 1).

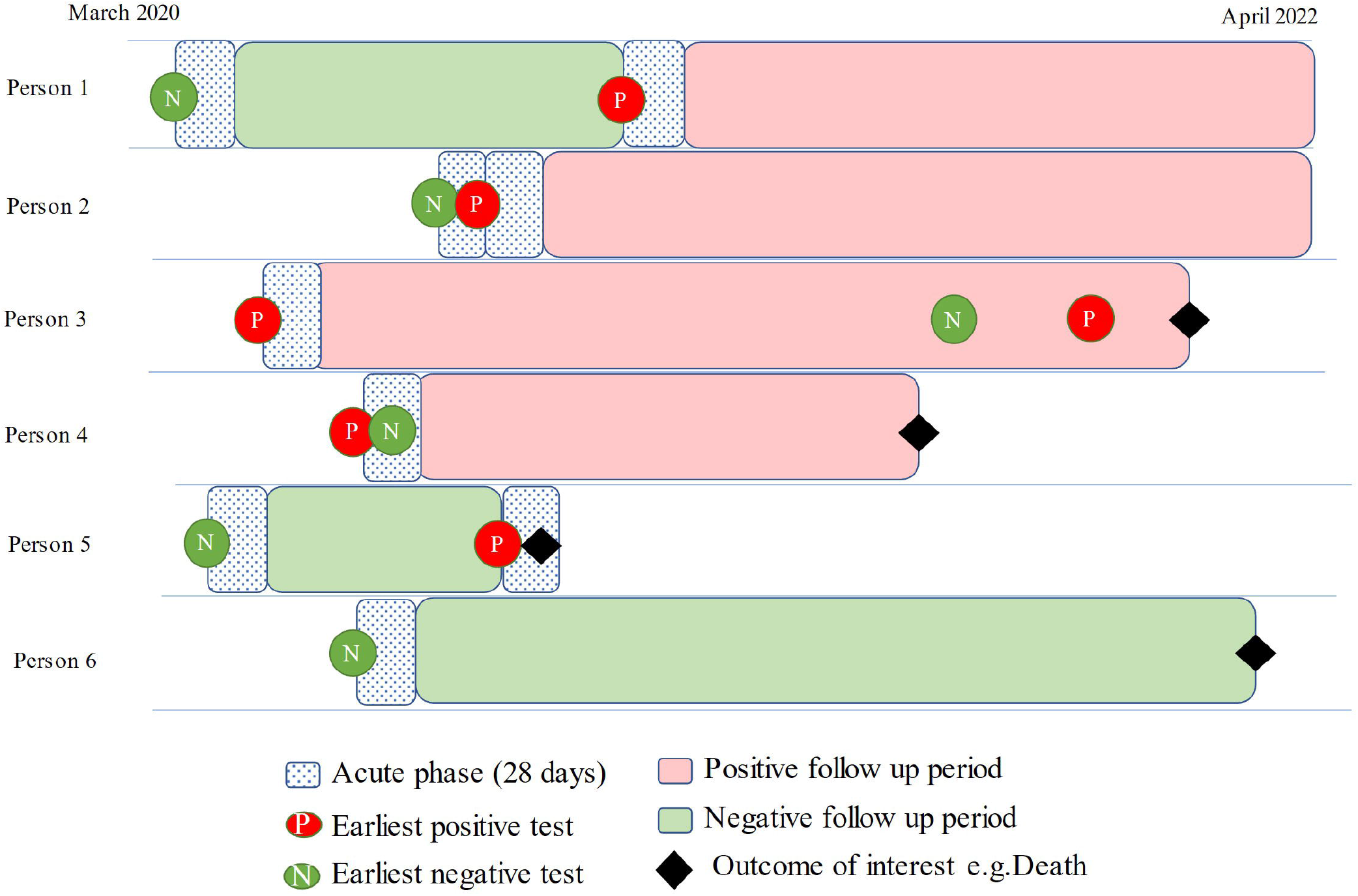

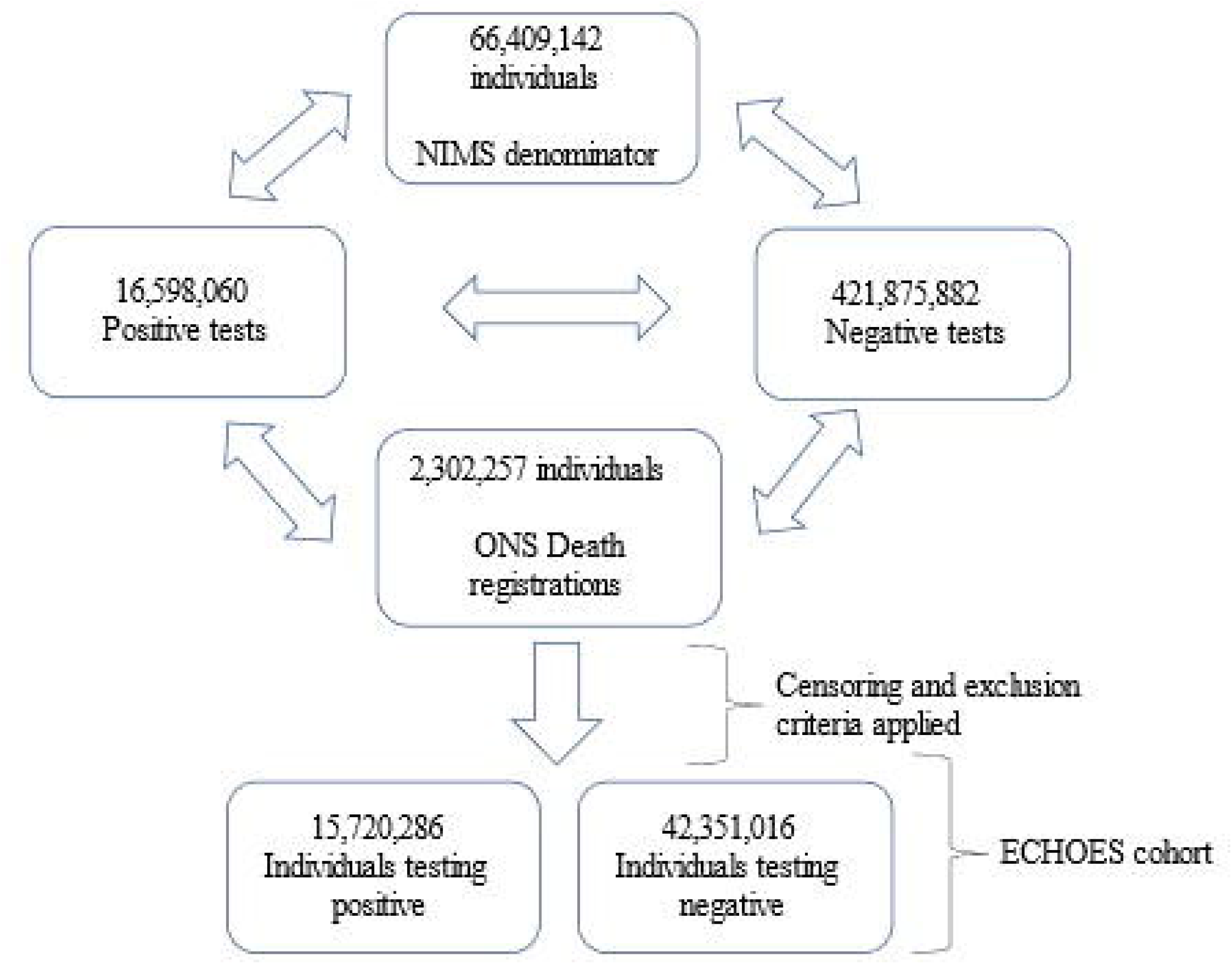

## Discussion

The development of ECHOES the dataset will facilitate the investigation of a range of post-acute health outcomes, including pre-specified health conditions and mortality. These investigations will allow greater understanding of how a SARS-CoV-2 infection episode is associated with changes in the risk of health outcomes, and the long-term impacts of COVID-19 more generally. Further, these data allow potential determinants of these outcomes to be accounted for in any analyses.

The inclusion of national level COVID-19 testing data in the ECHOES dataset creates a representative dataset, mirroring the overall population in England. The use of these data also allows the ECHOES dataset to capture those testing with asymptomatic infection and those with more severe disease testing in hospital settings.

A major strength of ECHOES is its large size. This facilitates the investigation of rare outcomes and for research questions unanswerable by smaller datasets to be answered. Using person level data from multiple data sources allows comprehensive linkage methods to be developed to create an accurate testing record for each individual. Furthermore, it facilitates the creation of a robust control group. This adds validity to the results produced, allows multiple study designs to be applied and means the findings are more generalisable to the general population.

Additionally, using multiple data sources means a range of health outcomes and risk factors can be investigated. Demographic factors such as age, sex, residential location, ethnicity and IMD can be considered, alongside vaccination status, comorbidities, pre-existing health conditions, acute disease severity and SARS-CoV-2 variant. Moreover, outcomes identified will not have to be considered independently and can be considered as indicators of future health outcomes.

Despite capturing all COVID-19 tests in England, certain individuals are excluded from the ECHOES dataset. Including only those with a valid NHS number may exclude specific population groups such as those who do not engage in healthcare. However, as ECHOES aims to compare outcomes between positive and negative groups rather than providing robust prevalence estimates for outcomes, this is unlikely to significantly impact the results produced.

Additionally, certain population groups that did not self-report their home test results may be excluded. How not being entitled to sick pay and other barriers to testing impacted different population groups needs to be considered when generalising results.

Early in the pandemic, prior to self-administered test availability, testing was limited to PCR tests in healthcare settings (March to June 2020) and non-hospitalised COVID-19 cases had limited access to testing. This may introduce inaccuracies in the results, however, these can by overcome by analysis by different time-periods.

Changes in test availability may lead to underrepresentation of population groups and misclassification of individuals. For example, individual’s earliest recorded positive tests could be a reinfection and individuals who tested negative may have also tested positive but never reported the result.

Lastly, the dataset is designed for long term follow-up individuals, with periodic linkage to updated health outcome data. As widescale community testing was ceased in England in April 2022 but COVID-19 has continued to circulate in the population, if individuals are infected with SARS-CoV-2 after this date, this may not be captured, therefore resulting in potential misclassification of the negative group over time, causing a reduction in risk difference between the two testing groups for certain outcomes. Furthermore, by not including individuals who’s first COVID-19 infection was after community testing was ceased may result in some under-estimation of the post-acute health outcomes of these of these later COVID-19 infections that may be distinct from those in the earlier time periods.

Despite these limitations, the ECHOES dataset offers a robust and comprehensive tool for investigating the post-acute health outcomes of COVID-19 in England. The dataset’s strengths will significantly enhance our understanding of the long-term impacts of SARS-CoV-2 infection.

## Data Availability

Any research conducted using the ECHOES dataset must be done as a formal collaboration with UKHSA. Proposals for collaboration will be considered by the Immunisation and Vaccine Preventable Diseases team to evaluate the scientific quality and feasibility of the proposal.

## Ethics, Caldicott and Data protection approvals

The production and analysis of this dataset has been approved by the UKHSA Caldicott Advisory Pane(15)l (Application reference CAP-2022-13) and the UKHSA Research & Public Health Practice Ethics and Governance Group (SUKHS REGG-Application reference NR0312). A Data Protection Impact Assessment was approved by DHSC Office of the Data Protection Officer in June 2023.

## Author Contributions

All authors were involved in the design of the ECHOES database and have contributed to the drafting of the manuscript. HA wrote the manuscript and lead on the design of the ECHOES database with oversight from ICM. KH led on the development of the database and development of entity resolution methods with support from CR and OP, APU, AJ, MCS have provided academic input into the design of the dataset. CC provided oversight specially for areas relating to vaccination data and GD and DPA have provided overall oversight.

## Acknowledgments

We thank the Data Operations team at UKHSA for coordinating data flows and the IT infrastructure to build the ECHOES dataset, the ECHOES steering group including Russell Hope, Guy Harling and Daniel Ayoubkhani and all colleagues working on COVID-19 Vaccine and Epidemiology at UKHSA who have inputted to discussions regarding the development of ECHOES.

## References

1. Zhang H, Zang C, Xu Z, Zhang Y, Xu J, Bian J, et al. Data-driven identification of post-acute SARS-CoV-2 infection subphenotypes. Nat Med [Internet]. 2023 Jan [cited 2023 Aug 14];29(1):226–35. Available from: https://www.nature.com/articles/s41591-022-02116-3

2. Symptom clusters in COVID-19: A potential clinical prediction tool from the COVID Symptom Study app. SCIENCE ADVANCES. 2021;

3. Pfaff ER, Girvin AT, Bennett TD, Bhatia A, Brooks IM, Deer RR, et al. Identifying who has long COVID in the USA: a machine learning approach using N3C data. The Lancet Digital Health [Internet]. 2022 Jul [cited 2023 Aug 14];4(7):e532–41. Available from: https://linkinghub.elsevier.com/retrieve/pii/S2589750022000486

4. UK Health Security Agency. Laboratory reporting to UKHSA: Guide for diagnostic laboratories [Internet]. 2022 Jan. Available from: https://assets.publishing.service.gov.uk/government/uploads/system/uploads/attachment_data/file/1108438/UKHSA_Laboratory_reporting_guidelines1_.pdf

5. NHS Digital [Internet]. [cited 2023 Mar 17]. Spine. Available from: https://digital.nhs.uk/services/spine

6. Twohig KA, Harman K, Zaidi A, Aliabadi S, Nash SG, Sinnathamby M, et al. Representativeness of whole-genome sequencing approaches in England: the importance for understanding inequalities associated with SARS-CoV-2 infection. Epidemiol Infect [Internet]. 2023 [cited 2024 Jul 26];151:e169. Available from: https://www.cambridge.org/core/product/identifier/S0950268823001541/type/journal_article

7. Covid-19 Unified Sample Dataset (USD).

8. GOV.UK [Internet]. [cited 2023 Mar 17]. Sources of surveillance data for influenza, COVID-19 and other respiratory viruses. Available from: https://www.gov.uk/government/publications/sources-of-surveillance-data-for-influenza-covid-19-and-other-respiratory-viruses/sources-of-surveillance-data-for-influenza-covid-19-and-other-respiratory-viruses

9. MikeRayMSFT. int, bigint, smallint, and tinyint (Transact-SQL) - SQL Server [Internet]. 2023 [cited 2023 Mar 24]. Available from: https://learn.microsoft.com/en-us/sql/t-sql/data-types/int-bigint-smallint-and-tinyint-transact-sql

10. rwestMSFT. LIKE (Transact-SQL) - SQL Server [Internet]. 2023 [cited 2023 Mar 17]. Available from: https://learn.microsoft.com/en-us/sql/t-sql/language-elements/like-transact-sql

11. DIFFERENCE (Transact-SQL) - SQL Server | Microsoft Learn [Internet]. [cited 2023 Mar 17]. Available from: https://learn.microsoft.com/en-us/sql/t-sql/functions/difference-transact-sql?view=sql-server-ver16

12. MikeRayMSFT. SOUNDEX (Transact-SQL) - SQL Server [Internet]. 2023 [cited 2023 Mar 17]. Available from: https://learn.microsoft.com/en-us/sql/t-sql/functions/soundex-transact-sql

13. Seghezzo G, Allen H, Griffiths C, Pooley J, Beardsmore L, Caul S, et al. Comparison of two COVID-19 mortality measures used during the pandemic response in England. International Journal of Epidemiology [Internet]. 2024 Feb 1 [cited 2024 Jul 26];53(1):dyad116. Available from: 10.1093/ije/dyad116/7249287

14. NCI Comorbidity Index Overview [Internet]. [cited 2023 Aug 10]. Available from: https://healthcaredelivery.cancer.gov/seermedicare/considerations/comorbidity.html

15. GOV.UK [Internet]. [cited 2023 Nov 9]. The Caldicott Principles. Available from: https://www.gov.uk/government/publications/the-caldicott-principles

