## Supplementary tables for "Evaluation of post-acute COVID-19 health outcomes (ECHOES) in England: The development of national surveillance system for long-term health outcomes following COVID-19"

Supplementary Table 1: Description of data sources included in the ECHOES dataset

| **Data source** | **Affiliated Organisation** | **Data provided** | **Primary uses** | **Key data provided** |
| --- | --- | --- | --- | --- |
| *Second generation surveillance system (SGSS)* | UK Health Security Agency | Positive COVID-19 testing data | COVID-19 data from SGSS are used to monitor trends in COVID-19 cases and as the basis for other areas of surveillance, such as monitoring genomic variations, residential clustering of cases and COVID-19 deaths. | surname and initial, date of birth and sex, ethnicity, residential postcode, hospital number and NHS number; reporting laboratory information, organism name, specimen type and specimen date |
| *Unified Sample Dataset (USD)* | UK Health Security Agency | Negative COVID-19 testing data | The USD is a repository for all COVID-19 testing data, at test level. COVID-19 data from USD are used to inform epidemiological analysis and modelling and to produce routine SARS -CoV-2 positivity metrics (11) | surname and forename, date of birth and sex, ethnicity, residential postcode, hospital number and NHS number; reporting laboratory information, organism name, specimen type and specimen date |
| *National immunisation management system (NIMS)* | UK Health Security Agency | COVID-19 Vaccination data | NIMS is used to monitor the roll out of vaccination programmes, including to assess vaccine coverage and effectiveness and to collect information on and assess any adverse reactions following vaccination in real time. | full name, date of birth and sex, ethnicity, residential postcode and NHS number; information on vaccination administered including location, date administered, manufacturer and batch information and an indicator for whether someone is classified as in a clinically vulnerable group. |
| *All-cause mortality data* | Office of National Statistics (ONS) | Death registration data | Using these data the ONS produce routine mortality statistics. | sex, usual address, place of birth, marital status, occupation, date of death. |
|  |  |  | Death registration data are also used for research and surveillance of disease. ONS provide UKHSA a weekly extract of all-cause mortality data for routine processing of mortality statistics and for outcome assessments of specific diseases such as COVID-19. | If investigation by the coroner is carried out, further details on the cause of death as determined by post-mortem are included (15). |
| *Hospital Episode Statistics data (HES)* | NHS England | Data on admissions, outpatient appointments and historical Accident and Emergency attendances at NHS hospitals | This dataset is used to collect national data on hospital activity to inform management and planning on NHS services (12,13) and is used in real time to inform severity of disease and disease associated hospitalisations | Person identifiers: NHS Number, age, sex, ethnicity, geographical information such as where patients were treated and their residential information. |
|  |  |  |  | Clinical information: diagnoses made during admission and procedures carried out. |
|  |  |  |  | Administrative information: date of admission and discharge, dates of diagnoses and treatment and dates of different episodes of care (17). |

Supplementary Table 2: Demographic breakdown of positive and negative individuals table

|  | **Positives ECHOES cohort** | **Positives ECHOES cohort %** | **Negatives ECHOES cohort** | **Negatives ECHOES cohort %** |
| --- | --- | --- | --- | --- |
| **Sex** | | | | |
| F | 8,405,774 | 53.66 | 20,434,603 | 52.28 |
| M | 7,260,177 | 46.34 | 18,648,664 | 47.71 |
| U | 245 | 0.00% | 651 | 0.00% |
| **Age band** | | | | |
| 00 - 04 | 357,547 | 2.28 | 1,658,961 | 4.24 |
| 05 - 09 | 1,020,639 | 6.51 | 1,962,771 | 5.02 |
| 10 - 14 | 1,408,129 | 8.99 | 2,798,206 | 7.16 |
| 15 - 19 | 1,153,530 | 7.36 | 2,506,354 | 6.41 |
| 20 - 24 | 1,202,897 | 7.68 | 2,551,885 | 6.53 |
| 25 - 29 | 1,311,232 | 8.37 | 2,802,910 | 7.17 |
| 30 - 34 | 1,361,732 | 8.69 | 2,963,247 | 7.58 |
| 35 - 39 | 1,321,581 | 8.44 | 2,792,477 | 7.14 |
| 40 - 44 | 1,272,340 | 8.12 | 2,558,679 | 6.55 |
| 45 - 49 | 1,138,503 | 7.27 | 2,536,607 | 6.491 |
| 50 - 54 | 1,077,414 | 6.88 | 2,679,591 | 6.86 |
| 55 - 59 | 913,195 | 5.83 | 2,564,410 | 6.56 |
| 60 - 64 | 663,148 | 4.23 | 2,147,049 | 5.49 |
| 65 - 69 | 431,676 | 2.76 | 1,744,598 | 4.46 |
| 70 - 74 | 350,372 | 2.24 | 1,694,643 | 4.34 |
| 75 - 79 | 252,461 | 1.617 | 1,256,426 | 3.21 |
| 79< | 429,800 | 2.74 | 1,864,835 | 4.77 |
| **NHS region** | | | | |
| East of England | 1,831,927 | 11.69 | 4,692,360 | 12.01 |
| London | 2,391,400 | 15.26 | 6,319,285 | 16.17 |
| Midlands | 2,925,347 | 18.67 | 7,087,851 | 18.13 |
| North East and Yorkshire | 2,448,185 | 15.63 | 5,578,286 | 14.27 |
| North West | 2,091,542 | 13.35 | 4,910,578 | 12.56 |
| South East | 2,482,974 | 15.85 | 6,478,795 | 16.58 |
| South West | 1,494,821 | 9.54 | 4,016,763 | 10.28 |
| **IMD decile** | | | | |
| 1 | 1,517,015 | 9.68 | 3,547,182 | 9.08 |
| 2 | 1,562,071 | 9.97 | 3,706,513 | 9.48 |
| 3 | 1,585,583 | 10.12 | 3,846,136 | 9.84 |
| 4 | 1,571,687 | 10.03 | 3,900,670 | 9.98 |
| 5 | 1,554,380 | 9.92 | 3,904,871 | 9.99 |
| 6 | 1,575,656 | 10.06 | 4,020,737 | 10.29 |
| 7 | 1,549,519 | 9.89 | 3,970,138 | 10.16 |
| 8 | 1,584,613 | 10.11 | 4,045,220 | 10.35 |
| 9 | 1,582,860 | 10.10 | 4,037,200 | 10.33 |
| 10 | 1,582,812 | 10.10 | 4,105,251 | 10.50 |
| **Vaccination status** | | | | |
| Unvaccinated | 8,460,152 | 54.00 | 30,964,379 | 79.23 |
| Fully Vaccinated | 7,206,044 | 46.00 | 8,119,539 | 20.77 |
